# Baseline correlation between pain, range of motion, disability, and health-related quality of life variables in subjects with frozen shoulder: A cross-sectional study

**DOI:** 10.1101/2024.07.29.24311165

**Authors:** A Poser, F Brindisino, D Feller, D Venturin

**Affiliations:** Physiotherapy private practice Kinè c/o Viale Venezia 13/Q San Vendemiano (Italy); Department of Medicine and Health Science “Vincenzo Tiberio”, University of Molise, Campobasso, Italy; Department of General Practice, Erasmus MC, University Medical Center, Rotterdam, The Netherlands; Department of Rehabilitation, Provincial Agency for Health Services of Trento, Trento (Italy)

## Abstract

Frozen shoulder (FS) is a debilitating condition affecting the glenohumeral joint. As FS frequently manifests as a chronic condition, it intensifies pain and leads to disability and to a deterioration in overall quality of life, affecting physical, behavioral, mental, and social dimensions.

While the impact of pain on disability is widely acknowledged, recent literature highlights a growing body of evidence indicating the correlation between pain and health-related, social, and mental distress and unfavorable outcomes in patients with FS.

Up to day, the need for pain to be avoided or alleviated as much as possible has been challenged, with a paradigm shift from traditional biomedical models of pain towards a biopsychosocial model of pain disorders. Research has further shown that psychological factors might affect the function and quality of life in patients with pain and can modulate the individuals’ pain experience and therefore may play a role in the development and/or maintenance of chronic pain states.

As the current healthcare pathway for subjects suffering from FS often inadequately addresses these healthcare needs, and professionals tend to predominantly concentrate on biological and clinical symptoms, the **aim** of this study will be to explore correlations among Patient-Reported Outcome Measures (PROMs) for pain, and disability, health-related domains, and ROM measurements in individuals with FS during their initial physiotherapy consultation

## BACKGROUND

Frozen shoulder (FS) is a debilitating condition affecting the glenohumeral joint, characterized by severe pain and progressive loss of both active and passive motion, ultimately resulting in unbearable pain and significant disability. Despite its wide prevalence, the precise pathological process of FS remains unclear, leading to uncertainty about its initial triggers and the underlying reasons for the pronounced mobility restrictions in the shoulder.

As FS frequently manifests as a chronic condition, it intensifies pain and leads to disability and to a deterioration in overall quality of life, affecting physical, behavioral, mental, and social dimensions. Moreover, FS foster emotions of frustration, uncertainty regarding prognosis, hindrance of social activities, and the forfeiture of professional and family roles. While the impact of pain on disability is widely acknowledged, recent literature highlights a growing body of evidence indicating the correlation between pain and health-related, social, and mental distress and unfavorable outcomes in patients with FS.

Furthemore, the need for pain to be avoided or alleviated as much as possible has been challenged, with a paradigm shift from traditional biomedical models of pain towards a biopsychosocial model of pain disorders. Research has further shown that psychological factors might affect the function and quality of life in patients with pain and can modulate the individuals’ pain experience and therefore may play a role in the development and/or maintenance of chronic pain states.

However, the current healthcare pathway for subjects suffering from FS often inadequately addresses these healthcare needs, as professionals tend to predominantly concentrate on biological and clinical symptoms.

In line with the patient-centered healthcare approach, it is vital to assess subjective health perceptions comprehensively in orthopedic subjects. It is equally important to investigate whether subjects’ self-perceived health aligns with objectively measured shoulder pain intensity, disability, and range of motion (ROM).

This study will **aim** to explore correlations among Patient-Reported Outcome Measures (PROMs) for pain, disability, health-related domains, and ROM measurements in individuals with FS during their initial physiotherapy consultation.

Understanding the distinct contribution of each element to the variability of perceived pain, can significantly influence clinical decision-making, enhancing the overall effectiveness of patient-centered healthcare practices.

### Aim of the study

The aim of this study is to investigate if ROM restriction and PROMs scores for disability and health-related domains have an impact on pain intensity in individuals with FS during their first physiotherapy consultation.

## METHODS

A cross-sectional study will be conducted.

The Strengthening the Reporting of Observational Studies in Epidemiology (STROBE) Statement for reporting will be used as a guideline for reporting.

This study will be asked to be approved from the Ethics Committee of University of Molise.

## INCLUSION CRITERIA

Subjects will be recruited from three private practice in Italy and will be screened for inclusion and exclusion criteria by a skilled physiotherapist with 10 years of experience in managing patients with shoulder pain.

Inclusion and exclusion criteria will be those from Kelley’s international guideline (i.e. ER with the arm at the side <50% compared to the contralateral side and ROM<25% in (at least) two or more other planes of movement. Symptoms had to be declared as stable or worsening for at least one month). Moreover, nontraumatic/micro traumatic onsed should be declared and a negative US examination (7.5-MHz probe US, Esaote, Genova, Italy) for excluding calcific tendinopathy and/or full thickness rotator cuff tear; and a negative x-ray evaluation for excluding gleno-humeral osteoarthritis (Grade 2 or more from Kellgren-Lawrence Classification)

All subject will be asked for a signed informed consent. The research procedures will adhered to the principles outlined in the Helsinki Declaration.

### Exclusion criteria

Patients that will not fully met the inclusion criteria will be excluded.

### Variables and measurement

*ROM measurement* will be collected from the all included studies for forward flexion and external rotatio at arm by side throught a digital inclinometer (JTECH Medical, Midvale, UT). Such measures will be performed using standardized procedures as descripted in the study from Poser et al., by skilled physiotherapists specialized in shoulder pathologies with more than ten years of experience, and blinded to all the other collected parameters. All values will be anonymously collected in an Excel file and de-identified. Only a chronological number will indicate each measure.

*Disability will be assessed througth the Disability of Arm, Shoulder and Hand (DASH) scale*. DASH questionnaire has 30 items rated on a 5-point Likert scale — 0 for no difficulty; 5 for impossible to perform — assessing challenges in activities which require upper limb function. The sum of each item is converted into a percentage, where a higher score reflected a greater disability. In this study, we will utilize the adapted and validated Italian version of the DASH questionnaire.

*Short Form Health Survey - SF-36*. SF-36 as administered to evaluate health-related quality of life. Specifically, it consists of 36 questions, divided into 8 health domains with 0-100 points for each domain, where higher values indicate a better health-related quality of life. In the present study, the subscores from the mental component summary will be considered, aiming to further assess the value of SF-36 mental health-related domains. The Italian-adapted version of SF-36 will be considered in this study.

*Pain by Shoulder Pain and Disability Index (SPADI) disability subscale*. The SPADI contains 13 items that assess two domains: a 5-item subscale that measures pain and an 8-item subscale that measures disability, ranging from 0 for no pain/disability to 10 for the highest pain/disability. In this study, an Italian-adapted and validated version of SPADI will be used, and only the sub-score for pain was considered.

## Data synthesis

The baseline data will be analyzed and a description of participants’ characteristics will be provided. Absolute and relative frequencies will be esteemed for categorical variables and mean and standard deviation (SD) for continuous variables.

The association of the SPADI-pain score as the dependent variable, with ROM in ER0°, FL, DASH, SF-36 vitality, emotional health, mental health, and social functioning will be calculated using a multivariable linear regression, which model all the independent variables simultaneously. The linear regression analysis will be adjusted with the following possible confounding variables: age, gender, dominant upper extremity, endocrine-metabolic pathologies (i.e. subjects suffering from diabetes and thyroid pathologies).

The assumption of absence of multicollinearity will be explored with the Variance Inflation Factor using a cut – off of 5. To compare the strength of the association of the independent variables with the dependent variable we will standardize the coefficients of the regression model.

From a strictly statistical point of view, there are no closed formulas to determine the sample size for fitting a multivariable linear regression model in an association study. Therefore, our sample size calculation will be based on the rule of thumb of at least ten participants per candidate coefficient. As we intended to use a regression model with twelve coefficients, we planned to recruit at least 150 patients. All the statistical analysis will be conducted with R in the R-studio software.

## RELEVANCE AND DISSEMINATION

The findings of this cross-sectional study review may increase clinicians’ knowledge about the association between pain and bio-psycho-social variables of interest, and consequently, may suggest a tailorised intervention in multiprofessional team.

On the whole, this study may add relevant information contributing to the improvement of the clinical management.

The results will be published in a peer-review journal and will be presented at relevant national and international scientific events.

## Data Availability

All data produced in the present work are contained in the manuscript

## AUTHORS’ CONTRIBUTIONS

All authors conceived, designed, drafted and approved the final protocol.

## COMPETING INTEREST STATEMENT

The authors declare no competing interest.

## FUNDING STATEMENT

This research will not receive any specific grant from funding agencies in the public, commercial, or not-for-profit sectors.

## Notes

### Competing Interest Statement

The authors have declared no competing interest.

### Funding Statement

This study did not receive any funding

### Author Declarations

Ethics Committee of University of Molise gave ethical approval for this work

